# Hospital length of stay throughout bed pathways and factors affecting this time: a non-concurrent cohort study of Colombia COVID-19 patients and an unCoVer network project

**DOI:** 10.1101/2022.11.17.22282466

**Authors:** Lina Marcela Ruiz Galvis, Carlos Andres Perez Aguirre, Juan Pablo Pérez Bedoya, Oscar Ignacio Mendoza Cardozo, Noël Christopher Barengo, Juan Pablo Sanchez Escudero, Jonathan Cardona Jimenez, Paula Andrea Diaz Valencia

**Author notes:** Corresponding author (LMRG). These authors contributed equally to this work (Lina Marcela Ruiz Galvis, Carlos Andres Perez Aguirre). These authors also contributed equally to this work (Juan Pablo Pérez Bedoya, Oscar Ignacio Mendoza Cardozo, Noël Christopher Barengo, Juan Pablo Sanchez Escudero, Jonathan Cardona Jimenez, Paula Andrea Diaz Valencia).

## Abstract

Predictions of hospital beds occupancy depends on hospital admission rates and the length of stay (LoS) according to bed type (hospital and intensive care unit beds). The objective of this study was to describe the LoS of COVID-19 hospital patients in Colombia during 2020-2021. Accelerated failure time models were used to estimate the LoS distribution according to each bed type and throughout each bed pathway. Acceleration factors and 95% confidence intervals were calculated to measure the effect on LoS of the outcome, sex, age, admission period during the epidemic (i.e., epidemic waves, peaks or valleys, and before/after vaccination period), and patients geographic origin. Most of the admitted COVID-19 patients occupied just hospital bed. Recovered patients spent more time in the hospital and intensive care unit than deceased patients. Men had longer LoS than women. In general, the LoS increased with age. Finally, the LoS varied along epidemic waves. It was lower in epidemic valleys than peaks, and became shorter after vaccinations began in Colombia than before. Our study highlights the necessity of analyzing local data on hospital admission rates and LoS to design strategies to prioritize hospital beds resources during the current and future pandemics.

## Introduction

SARS-CoV-2 is difficult to be eradicated, because of its ubiquity [1], the incomplete vaccine coverage [2,3], and virus evolution [4]. Hence, ongoing strategies to deal with the endemic presence of SARS-CoV-2 in populations over the long term will be needed [3]. It is important to understand the impact of COVID-19 on hospital capacity and to be prepared for hospital bed demand by COVID-19 patients [5]. High demand for beds could severely decrease the quality of services in part because of the limited number of regular beds and Intensive Care Units (ICU) [6]. As a consequence, designing strategies to prioritize hospital beds resources is mandatory to cope with new waves of the epidemic or endemic outbreaks [7].

One of the main indicators needed for any predictions is the estimation of the number of hospital beds occupancy, which is essential in managing the resources allocation [8]. This indicator is a function of hospital admissions rates and the total LoS in particular bed types (hospital and Intensive Care Unit (ICU) beds) [7,8]. It is possible to model the rate of hospitalization in many settings based on estimated epidemic curves [7]. On the other hand, statistical models allow us to estimate the LoS through the observation of patients’ bed pathways (BPs), which is the sequence of transfers between bed types during a hospital stay [6,9]. Additionally, different factors such as sex, age, comorbidities, and the variations in the treatments, medical staff, equipment, and access to health services could potentially explain differences in the hospital LoS and the level of care needed [9,10]. However, up-to-date, there is little scientific information available on the characteristics of hospital LoS of COVID-19 patients in Colombia. Therefore, the objective of this study was to describe the hospital LoS of Colombia COVID-19 patients during 2020-2021.

## Methodology

### Database description

Publicly available data of all registered COVID-19 hospital patients from March 15^th^ 2020 to August 17^th^ 2021 were retrieved from the Colombia National Health Institute [11]. The diagnosis of SARS-CoV-2 was confirmed in all patients by diagnostic tests [12]. The database included in-hospital patients who either recovered from Covid 19 or deceased. Recovered patients were those with a negative test result and/or with 21 days without symptoms since the symptoms onset [12]. This study was developed within the unCoVer Horizon 2020 project framework [13].

This study followed the Good Clinical Practice guidelines and the guidelines of the Helsinki Declaration. This study was a secondary data analysis using de-identified data downloaded from the Colombian Institute of Public Health that is publicly available [11]. Ethical approval was waived by IRB of the Faculty of Public Health, Universidad de Antioquia since the analysis was considered nonhuman subject research.

### LoS by bed pathway and stage

Daily hospital bed ubication by patient was retrieved and ordered chronologically to identify all possible stages (i.e., bed locations) and BPs. For each patient the time of stay in each stage of its BP was estimated [6]. The LoS of the hospitalization until the first discharge was included in the analysis. Taking into account the maximum times previously reported in the literature [6,8–10], a right truncation of the data was made to restrict the maximum length of hospital stay to 100 days.

An Accelerated Failure Time (AFT) model was run for each stage of every BP with different distributions (i.e., Lognormal, Gamma, and Weibull) [14,15]. AFT model was used because the aim was to estimate the times to failure and the effect on time of some covariates. This is a parametric model in the statistical field of survival analysis that offers an alternative to the often used Proportional Hazard models [15]. For this survival analysis, the variable of interest was the amount of time measured in days that a patient occupies a type of bed. The Akaike Information Criterion (AIC) was used to select the best fitting parametric distribution. For this survival analysis we used RStudio 2021.09.0.

We used the estimated parameters of each best fitting distribution for each stage in each BP to estimate the median of the LoS distribution and its 95% confidence interval [6]. For this, we implemented a Monte Carlo method (S1 Fig).

**Fig 1.**
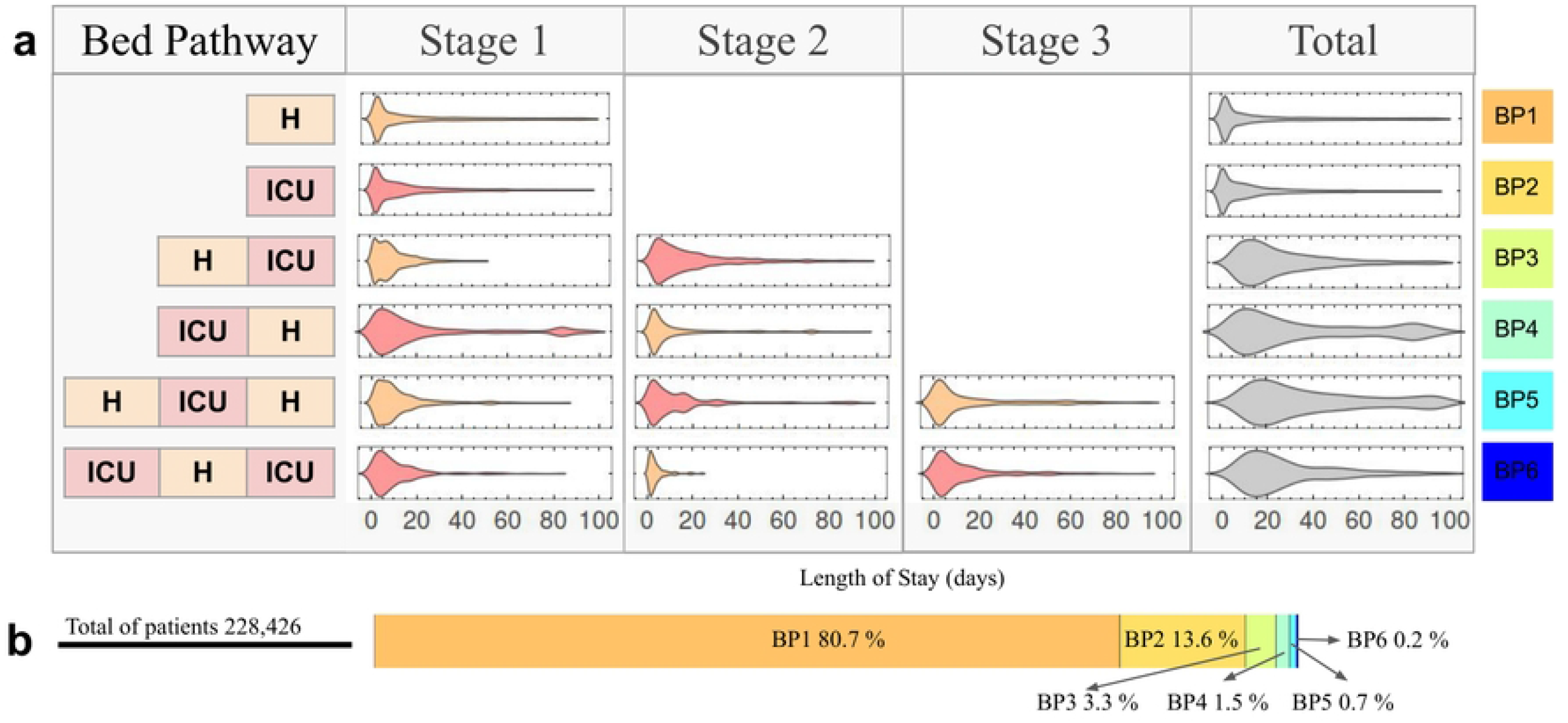
Bed pathways of Colombia COVID-19 patients. (**a**) For each bed pathway (H - [BP1]; ICU - [BP2]; H, ICU - [BP3]; ICU, H - [BP4]; H, ICU, H - [BP5]; ICU, H, ICU - [BP6]; according to bed types: H: Hospital (light orange color), ICU: Intensive Care Unit (pink color) is shown the LoS distribution for each stage or bed types and the total of bed pathway (in gray color). (**b**) The proportions of hospital admissions entering each bed pathway.

### Association between LoS and covariates

The four most common BPs were chosen in this study to estimate the effect of covariates on the LoS in each stage of each BP. For each covariate, an AFT model was run for each stage of each bed pathway. The AFT model estimates the accelerated factor (*θ*) which allows to evaluate the effect of predictor variables on survival time just as the hazard ratio allows the evaluation of predictor variables on the hazard [15]. The covariates were BP, outcome (i.e., recovery or death), age (i.e., 1-25, 26-50, 51-75, and >75 years), sex (men *vs* women), and the epidemic phase in which the patient was admitted and discharge/death as explained below.

We identified some phases of the COVID-19 epidemic curve as explained in the SI. These phases are characterized in epidemiology as waves, and peaks or valleys as is detailed in S2d and S3d Figs, respectively. Then, each patient was classified according to a specific wave, and peak or valley depending on the dates in which they were admitted and discharged from the hospital or died. The study population was also categorized into patients admitted before and after the vaccination period in Colombia, which started in February 2021 [16]. Additionally, an AFT model was used adjusted for all mentioned covariates.

Finally, an AFT model for the two most common BPs was calculated to estimate the association between the LoS and the geographic region of origin of each patient. The median of LoS for the best fitting distribution was estimated as previously described. Then, a k-means clustering algorithm was applied to group the geographical regions with similar medians.

All the code for statistical analyses and databases used in this study can be accessed at GitHub. The processing of raw data to get the databases used are in GitHub. All authors had full access to the study data and accepted the responsibility to submit for publication.

## Results

### LoS by bed type and bed pathway

In total, 245,371 COVID-19 patients were hospitalized, they were recovered or deceased between March 15^th^ 2020 and August 17^th^ 2021 in Colombia. They represented 5.03 % of the total number of COVID-19 cases in Colombia on August 17^th^ 2021. After identifying their BP and filtering those with LoS lower than 100 days, 228,426 COVID-19 patients remained to be included for the data analysis (Fig 1). The COVID-19 patients in Colombia could occupy two types of bed: a hospital or ICU bed. Six different BPs were possible. We identified that the BPs have between one to three stages which represent the different bed transfers of a patient during its hospital stay, and noticed that BP1 or BP2 only have one stage (stage 1), but for the BP5 or BP6 the patients have three stages (stage 1 to 3) (Fig 1). Each one of the LoS distributions were best fitted to the Lognormal distribution except the stage 1 and 2 of BP3 and the total distribution of BP4, these were best fitted to Gamma distribution.

Table 1 presents the median and IQR of the length of stay in each stage of each bed pathway. Most of the patients only occupied a hospital bed (80.70%) or an ICU bed (13.58%), with median times of 6.5 (95% CI 6.2-6.7) and 6.7 (95% CI 6.5-6.9) days, respectively. The remaining patients occupied both hospital and ICU beds at different moments (5.7%).

**Table 1.** Median and IQR of the length of stay in each stage of each bed pathway. These were estimated with the time distributions that best fitted according to the AFT models.

| Bed Pathway | n | % | Stage 1 |  | Stage 2 |  | Stage 3 |  | Total |  |
| --- | --- | --- | --- | --- | --- | --- | --- | --- | --- | --- |
|  |  |  | x | IQR | x | IQR | x | IQR | x | IQR |
| **H (BP1)** | 184,340 | 80.70 | 6.48<br>(6.23-6.75) | 2.16 -<br>19.45 | N/A | N/A | N/A | N/A | 6.49<br>(6.23-6.75) | 2.16 -<br>19.45 |
| **ICU (BP2)** | 31,032 | 13.58 | 6.69<br>(6.46-6.94) | 2.56 -<br>17.53 | N/A | N/A | N/A | N/A | 6.69<br>(6.46-6.94) | 2.56 -<br>17.53 |
| **H,ICU (BP3)** | 7,632 | 3.34 | 7.24<br>(7.07-7.43) | 3.44 -<br>13.27 | 12.38<br>(12.04-12.72) | 5.23 -<br>24.45 | N/A | N/A | 20.57<br>(20.17-20.97) | 12.16 -<br>34.77 |
| **ICU,H (BP4)** | 3,341 | 1.46 | 10.68<br>(10.34-11.03) | 4.41 -<br>25.90 | 4.92<br>(4.75-5.08) | 1.95 -<br>12.46 | N/A | N/A | 26.98<br>(26.36-27.62) | 13.19 -<br>48.40 |
| **H,ICU,H (BP5)** | 1,653 | 0.72 | 6.77<br>(6.59-6.95) | 3.30 -<br>13.86 | 6.73<br>(6.53-6.94) | 2.86 -<br>15.82 | 5.79<br>(5.60-5.99) | 2.30 -<br>14.58 | 30.54<br>(29.99-31.08) | 18.68 -<br>49.90 |
| **CU,H,ICU (BP6)** | 428 | 0.19 | 6.47<br>(6.30-6.64) | 3.12 -<br>13.39 | 2.68 (2.62-2.74) | 1.45 -<br>4.92 | 6.37 (6.18-6.57) | 2.76 -<br>14.70 | 21.82 (21.44-22.21) | 13.38 -<br>35.59 |
| **Total** | 228,426 | 100 |  |  |  |  |  |  |  |  |

H - [BP1]; ICU - [BP2]; H, ICU - [BP3]; ICU, H - [BP4]; H, ICU, H - [BP5]; ICU, H, ICU - [BP6]; according to bed types: H: Hospital, ICU: Intensive Care UnitH Hospital bed; n: number of hospital admissions entering each bed pathway, %: proportion of hospital admissions entering each bed pathway, $\bar{x}$ : Median and 95% C.I., IQR Interquartile range. N/A non available data.

### Association between LoS and some covariates

Table 2 presents the acceleration factors of the LoS for each stage of the four most common BPs, and S1 Table presents the times or LoS for each stage of the two most common BPs. The LoS of each bed type depended on the BP. For instance, patients in BP3 and BP4 spent 0.95 (*θ*, 95% C.I 0.91-0.98) and 0.76 (*θ*, 95% C.I 0.72-0.80) times less time in hospital bed than those in BP1 (Table 2). They have a hospital bed LoS of 6.16 (*θ*, 95% C.I 5.92-6.41) and 4.92 (*θ*, 95% C.I 4.73-5.12) days, respectively (S1 Table). In contrast, patients in BP3 and BP4 spent 1.51 (*θ*, 95% C.I 1.46-1.56) and 1.60 (*θ*, 95% C.I 1.52-1.68) times more time in ICU beds than those in BP2.

**Table 2.**
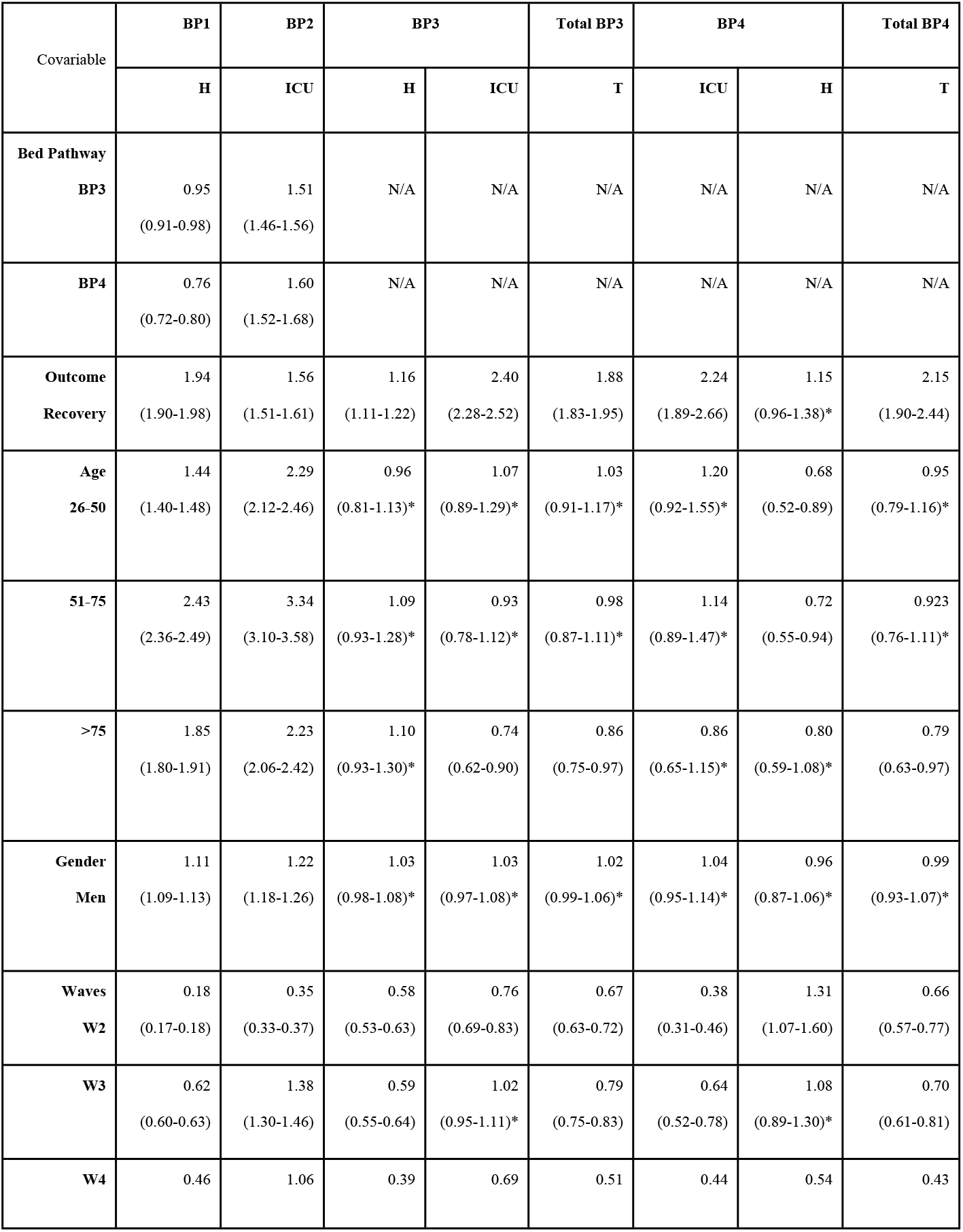
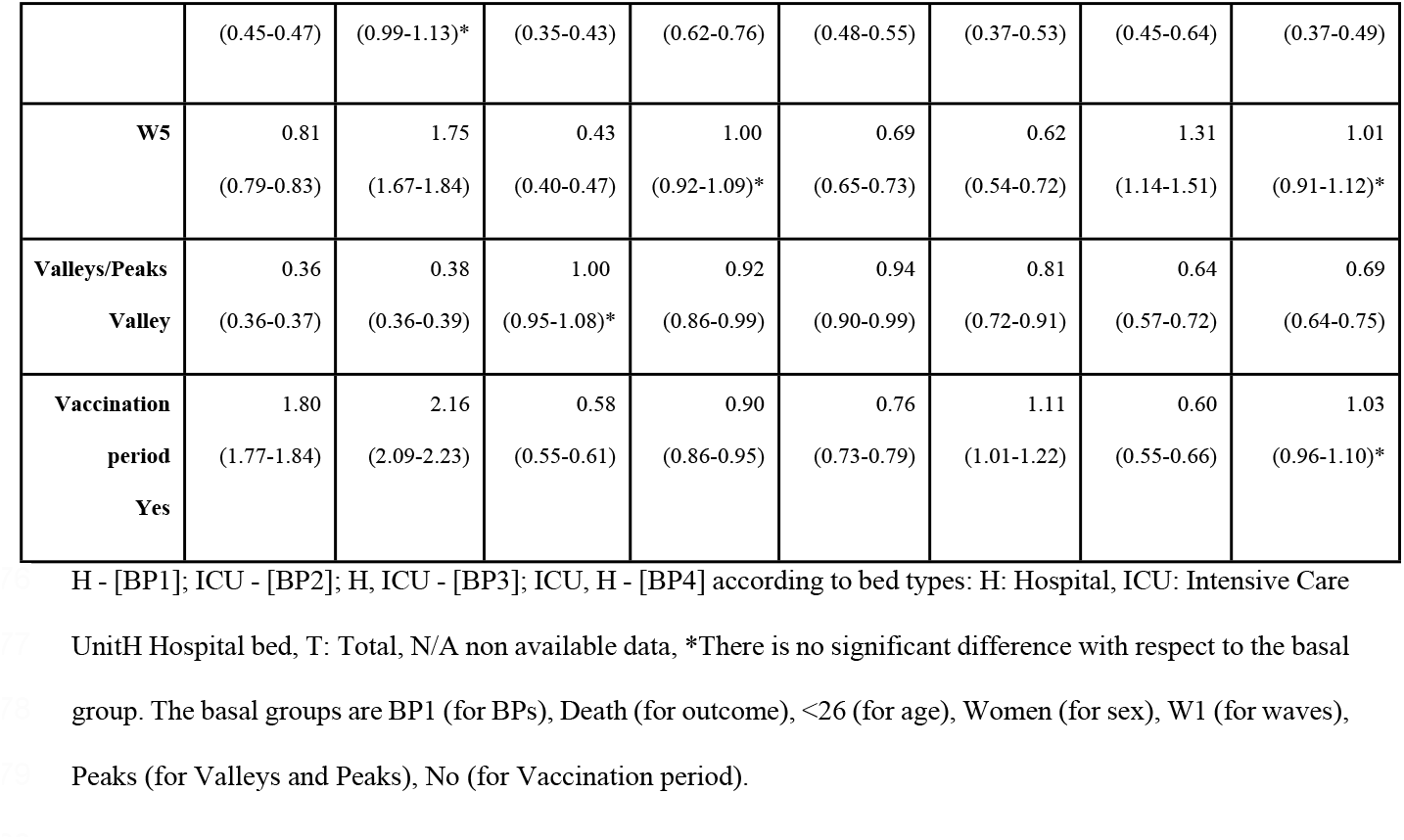
Acceleration factors of the length of stay in each bed type and each bed pathway. Those were calculated in different AFT models for bed pathway, outcome, age, gender, waves (W), peaks and valleys, and vaccination period.

The LoS in each bed type and BP was significantly higher for recovered patients than for deceased ones, except for the hospital bed (stage 2) in BP4. Age had a significant effect in the LoS mainly for BP1 and BP2. In general for both pathways, the LoS in the hospital and ICU increased with age, but that increase was non-monotonic. Moreover, sex was associated with the LoS only for BP1 and BP2. Men had 1.11 (*θ*, 95% C.I 1.09-1.98) and 1.22 (95% C.I 1.18-1.26) times the LoS in hospital and ICU beds compared with women.

The LoS were statistically significantly different throughout the epidemic COVID-19 waves compared with the first wave in almost all BPs. The LoS in hospital for the BP1 was lower in W2, W3, W4 and W5 than in W1 (i.e., *θ* 0.18, 95% C.I 0.17-0.18; *θ* 0.62, 95% C.I 0.60-0.63; *θ* 0.46, 95% C.I 0.45-0.47; and *θ* 0.81, 95% C.I 0.79-0.83, respectively), with the maximal diminution for W2. In contrast, the LoS in ICU for the BP2 was higher in W3 to W5 with respect to W1, only with a diminution of 0.35 (*θ*, 95% C.I 0.33-0.37) in W2. The LoS for BPs was lower in valleys than peaks.

For BP1 and BP2, the LoS was larger in the vaccination period than before it (*θ* 1.80, 95% C.I 1.77-1.84; *θ*, 2.16, 95% C.I 2.09-2.23, respectively). Thus, the majority of patients occupying a hospital or ICU bed spent more time in these beds after vaccination in the population began than before. Finally, the association factor of the effect of the admission period during waves and vaccination changed when the model is adjusted for all the covariates (S2 Table).

### Association between LoS and geographical region of origen

Figs 2a and 2b present the cluster analysis for patients’ geographic region of origin according to the median of the LoS in hospital and ICU bed for BP1 and BP2. There were five and four clustering for hospital and ICU beds.

**Fig 2.**
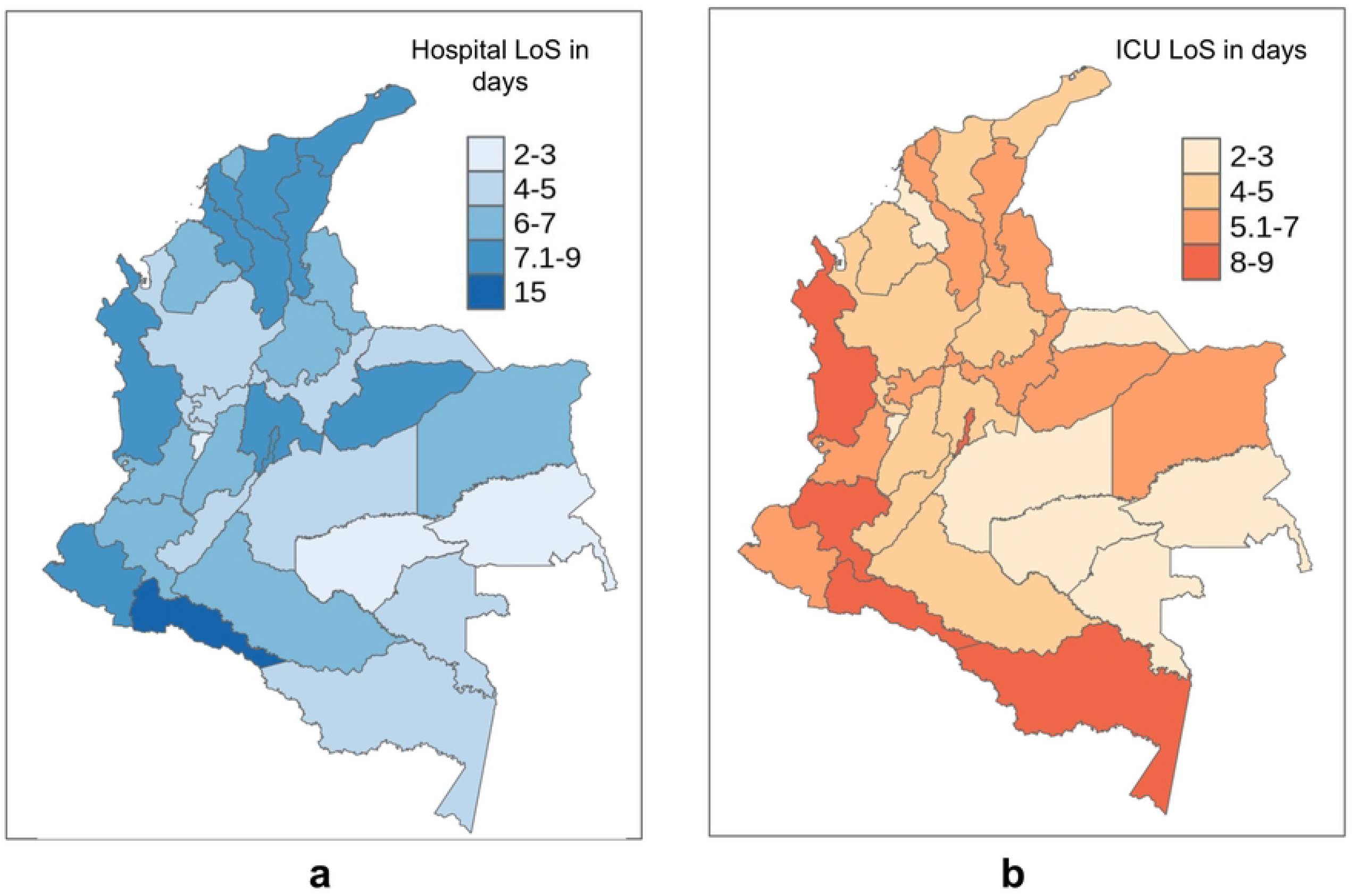
Clustering of patients regions of origin according to similarity in the median of LoS. (**a**). in hospital bed for bed pathway 1 (BP1), (**b**). in ICU bed for bed pathway 2 (BP2).

## Discussion

The Colombia COVID-19 patients occupied mainly a hospital or ICU bed during the hospitalization between March 2020 to August 2021. The length of stay in each one of these beds dependeds on the outcome (i.e., recovery or death), age, sex, the epidemic period in which the patient gets admitted and discharge/death (i.e., epidemic waves, peaks/valleys, and before/after the vaccination period), and the geographical region of origen. For instance, recovered patients spent more time in the hospital and ICU than deceased patients. Men had larger LoS for both hospital and ICU beds than women. In general, the LoS in the hospital and ICU increased with age. Finally, the LoS in both hospital and ICU beds varied along epidemic waves. It was lower in epidemic valleys than peaks, and was shorter after vaccinations began in Colombia than before.

### LoS by bed type and bed pathway

The bed types and bed pathways were similar to those reported in previous studies [6,8]. Our data also revealed that the majority of patients occupied a hospital bed without entering into an ICU, similar to hospital patients in England [6]. This may be due to the low severity of COVID-19 [5]. The majority of the LoS distributions in each bed type and each bed pathway were well fitted to log-normal distributions. As in previous studies, the LoS distributions were right-skewed due to a minority of patients with long hospital stays [6,8,10,17]. The tail of the distribution should not be ignored since these few patients can block beds for a long time and form a heavy burden on capacity [9].

Both the median and IQR of hospital and ICU LoS for Colombia patients are similar to the previously reported. However, our IQRs had an upper limit higher than those reported previously. For instance, it was reported that the median (IQR) of the total hospital stay is 14 (10–19) days in China, whereas it was 5 (3–9) days outside of China [9]. A study in New York reported comparable estimates for total LoS (i.e., median 4.5, IQR 2.4–8.1) [18]. Studies from Northern Italy [18,19], Japan [20], and California and Washington [21] reported longer estimates of LoS than our data. On the other hand, the median (and IQR) of the ICU stay is 8 (5–13) for China and 7 (4–11) outside of China [9,22]. In England, the mean ICU length of stay is 12.4 days [8].

### Association between LoS and covariables

We showed that recovery patients spent more time in hospital or ICU beds than deceased patients. This was also found in China [9], and Turkey [5]; different results were found for California and Washington patients with a hospital stay median of 9.3 and 12.17 for survivors and non-survivors, respectively [21]. Also different results were found for London in ICU occupancy [23]. However, some argue that the outcome does not have practical implications to estimate the bed occupancy and influence decision-making [9].

Our data found that older people spend more time in hospital and ICU beds than young people, similar results were also reported in previous studies in hospitals in England and Turkey [5,8]. Also similar to previous studies, men stay longer in hospital and ICU beds than women [5,17]. Some studies concluded that the effect of sex was small and non-significant [8,24].

The hospital LoS decreased during the epidemiological waves, but the ICU LoS increased. Studies in South Africa, Belgium, England, US and Canada also concluded that the LoS in hospitals decrease throughout the time (i.e., waves or weeks) [8,10,17,25]. Contrary to our results, previous studies in England, US, and Canada concluded that the LoS in ICU also decreases throughout time [8,17,26].

However, none of these studies had an explanation for this tendency. Here, we identified that both the number of admitted patients and the age distribution of patients throughout the waves could possibly affect the LoS (S4 Fig). For instance, the LoS increased/decreased with the increase/decrease in the number of hospital admissions and the increase/decrease in the median age of the admitted patients. We also hypothesized that the decrease in hospital LoS as reported in other studies might be related to improved clinical experience and improved treatments over the course of the epidemic [10,17,27]. However, there was an increment in the hospital LoS when these times were adjusted by all the covariates.

The hospital and ICU LoS were lower in valleys than in peaks. On the other hand, these LoS were higher in the vaccination period than before it; but when these times are adjusted by all covariates, these LoS were lower in the vaccination period than before it. Finally, we found different LoS for patients coming from different geographical regions in Colombia. As previously mentioned, this could be due to different patients profiles (i.e. percentage of comorbidities, ages group, sex), treatments, and general quality of the health care system.

## Contributions

Although LoS is an important factor in predicting the needs of hospital resources, it was rarely a primary outcome in previous studies [17]. Most of the studies estimating LoS were derived from China [9], and they reported the LoS for total hospital and ICU stay without describing the bed pathway [9].

To the best of our knowledge, there are no previous studies of the LoS in Colombia. While there are some similarities with previous studies in the LoS and factors associated with them, there are also some differences. This discrepancy is due to countries differing in the characteristics of factors associated with LoS [17]. For instance, countries differ in their demographics characteristics, in the management of the pandemic, and have different COVID-19 care guidelines (i.e., the criteria for hospital admission and release might be distinct in different countries)[9,10].

On the other hand, any prediction model of bed occupancy is sensitive to the value and distribution being assumed for LoS, with strong implications for policy and planning [9]. Therefore, there is a need for local information on LoS for COVID-19 patients [17]. This is important to set with appropriate LoS values the local models that predict the number of beds required and then to provide accurate predictions [17,28].

## Limitations

This study is based on a secondary analysis of publicly available data that limits the availability of data of what was collected during the pandemic in Colombia. Also, due to the nature of the data, information or selection bias such as missed information or misclassification of the BPs is possible. Finally, there are other factors that may be considered as potential confounders affecting the LoS that were not included in the analysis, such as patient comorbidities, other health conditions or social determinants of health that were not available in the public database.

## Conclusion

While our results share some similarities with previous studies in other countries, the differences in the LoS and the factors affecting the LoS highlight the necessity of local information to parameterize models and improve the predictions of the hospital bed occupancy in Colombia. Future studies might consider comorbidities information of the patients. Finally, these findings help to prepare for risk prevention and control in advance according to the local demographics features and the current situations of medical resources.

## Data Availability

All the code for statistical analysis and databases used in this study can be accessed at https://github.com/LinaMRuizG/unCoVer_ColombiaLoSCOVID-19. The processing of raw data to get the databases used are in https://github.com/LinaMRuizG/unCoVer_processingData. All authors had full access to the study data and accepted the responsibility to submit it for publication.

https://github.com/LinaMRuizG/unCoVer_processingData

## Acknowledgements

We thanks the ReFReCA project which is financed by the Ministry of Science, Technology and Innovation of Colombia - Minciencias (code 111584467754). We also thanks the Physics Fundamentals and Teaching Group (FEnFisDi) at the Physics Institute of the University of Antioquia for let us use their server to run some simulations.

## Supporting information

**S1 Fig.**
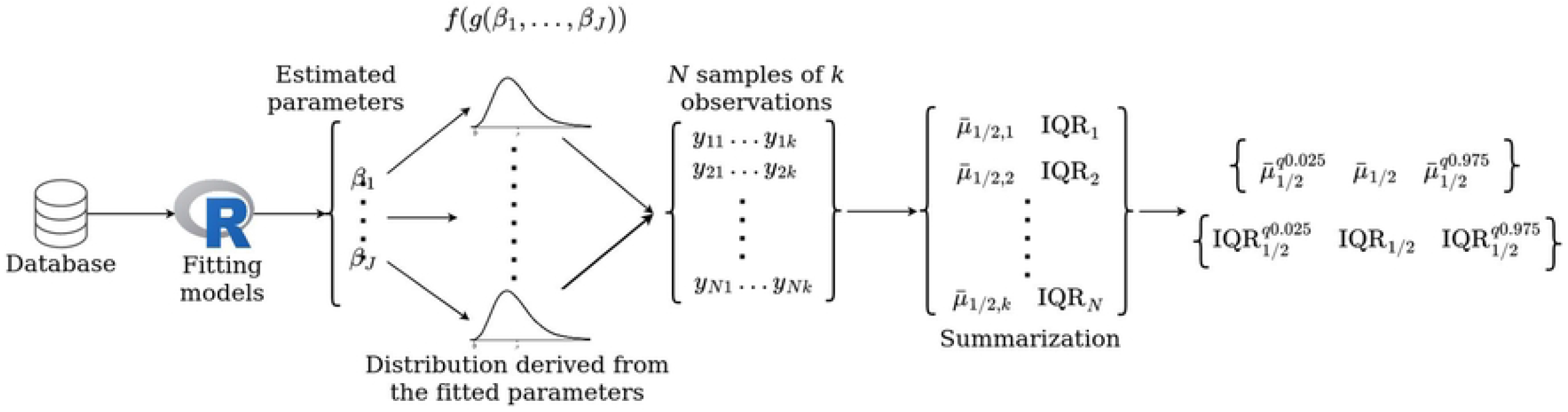
Estimation of the median, IQR, and their 95% confidence interval of each AFT model.

**S2 Fig.**
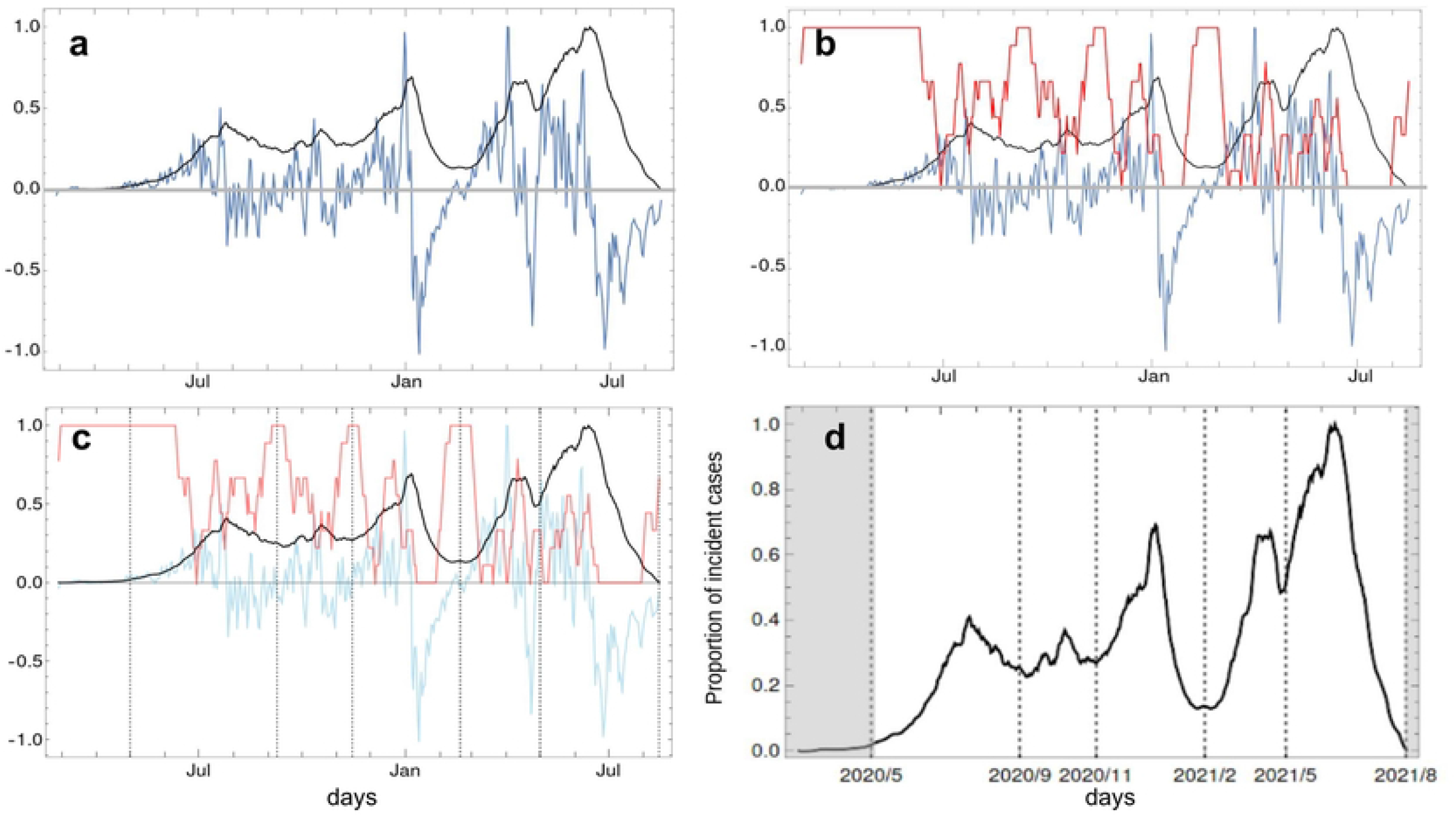
Estimation of waves of the epidemic curve of SARS-CoV-2 daily infected cases in Colombia.

**S3 Fig.**
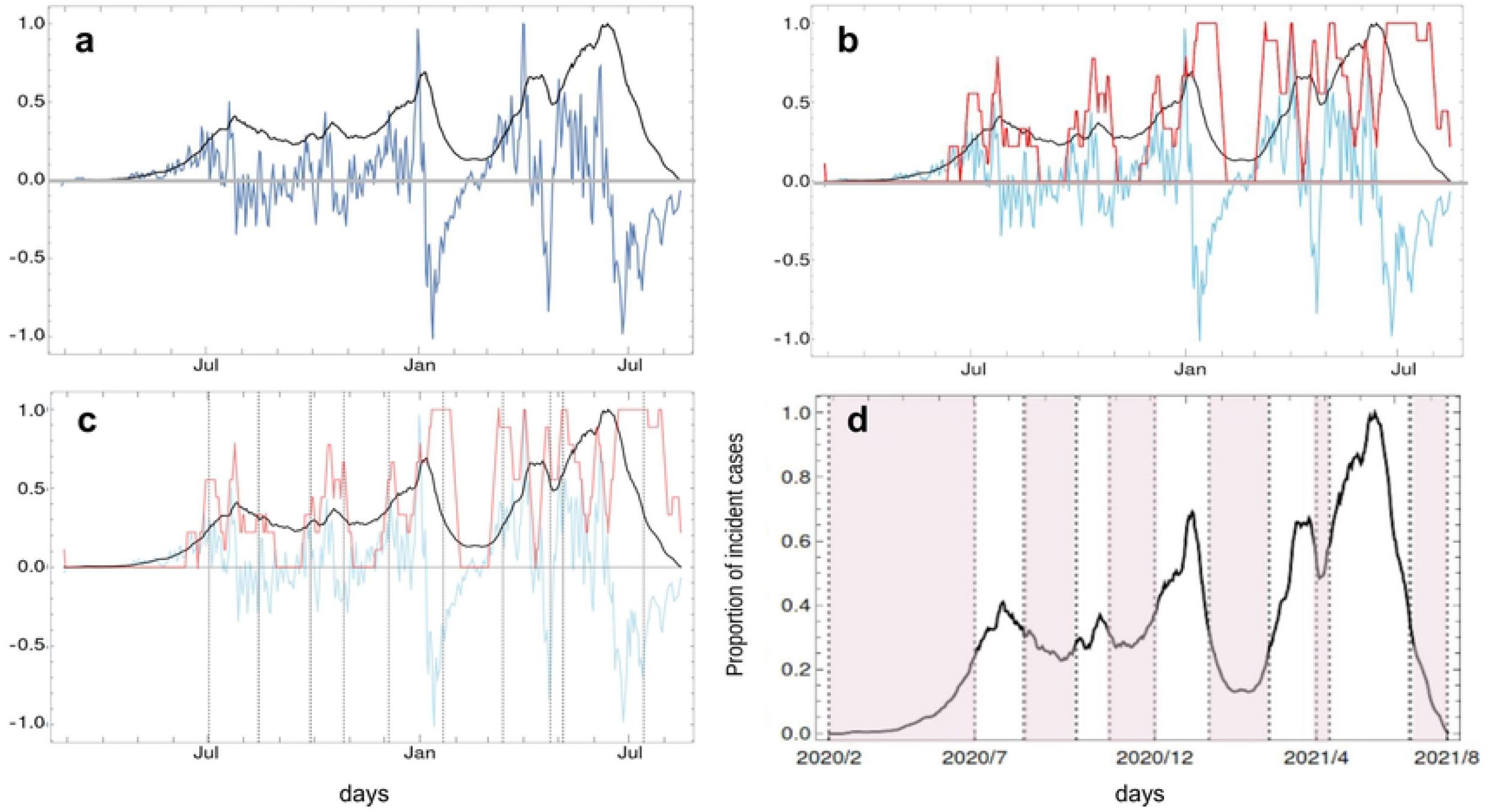
Estimation of peaks and valleys of the epidemic curve of SARS-CoV-2 daily infected cases in Colombia.

**S1 Table. Length of stay for Hospital (BP1) and ICU (BP2) pathways according to covariables**. Those were calculated in different AFT models for bed pathway, outcome, age, sex, waves, peaks and valleys, and vaccination period. We get the lognormal distributions parameters from each AFT model and performance sampling to estimate the median and IQR.

**S2 Table. Acceleration factors of the length of stay in each bed type and each bed pathway for a model adjusted by all covariable**.

**S3 Table. Median and IQR of the length of stay in hospital (BP1) and ICU (BP2)**. We get the Lognormal distributions parameters from each AFT model and performance sampling to estimate the median and IQR.

**S4 Fig.**
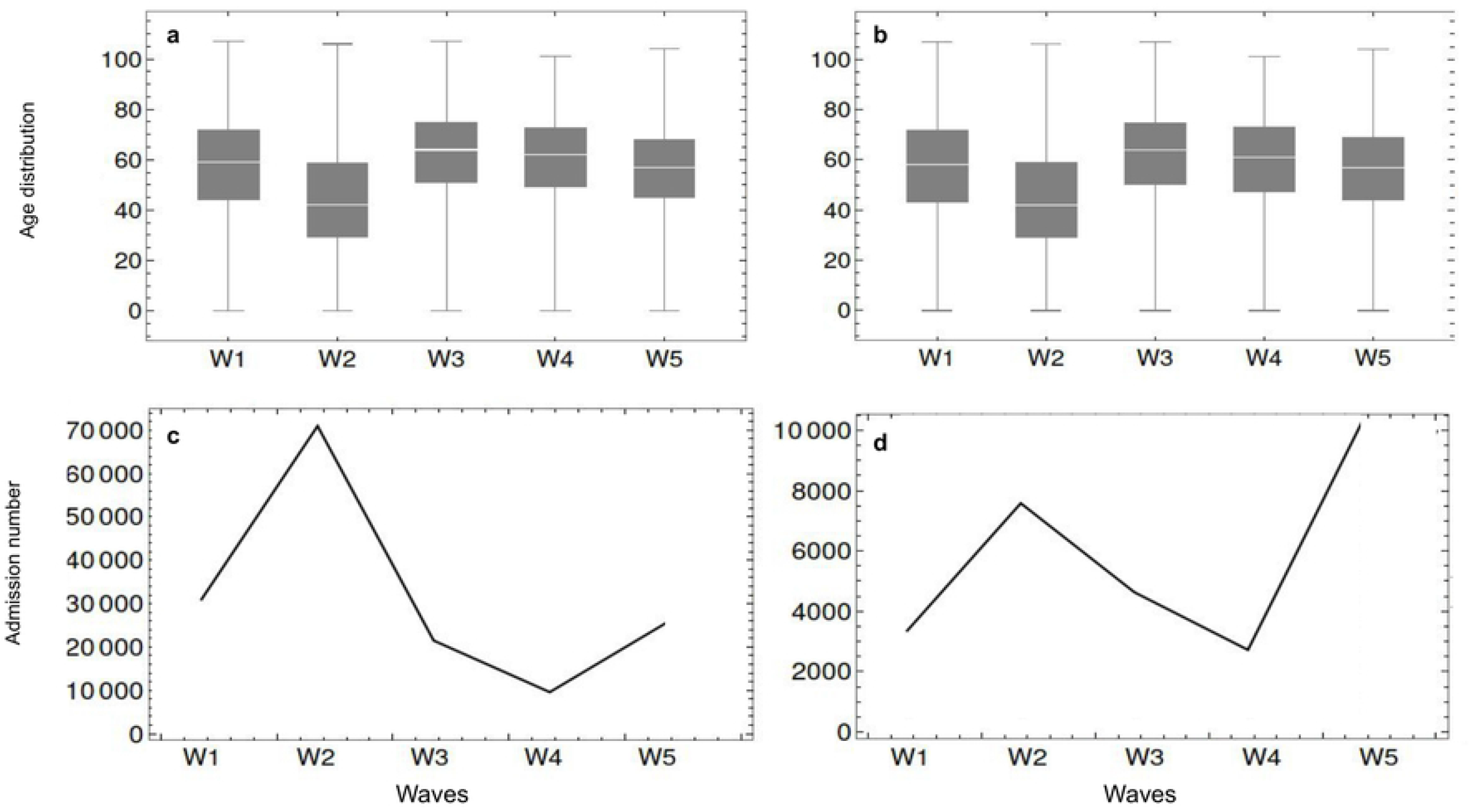
Age distribution and admission number throughout the waves. (a). Age distribution of patients with BP1 (only hospital bed), (b). Age distribution of patients with BP2 (only ICU bed), (c). Admission number of patients with BP1 (only hospital bed), (d). Admission number of patients with BP2 (only ICU bed).

## Notes

### Competing Interest Statement

The authors have declared no competing interest.

### Funding Statement

This project was funded by the European Union’s Horizon 2020 Research and Innovation Program (Grant Agreement No 101016216). The funders do not played any roles in the study design, data collection and analysis, decision to publish, and preparation of the manuscript

